# The importance of timing of a population level intervention on COVID-19 mortality

**DOI:** 10.1101/2020.04.19.20071845

**Authors:** Jasper S. Johnston, Eloïse S. Johnston, Sebastian L. Johnston

## Abstract

COVID-19 poses an immense and immediate threat to global public health. Population level interventions (PLIs) impact this threat, with estimable large effects on reducing mortality. Many countries worldwide have currently zero/low mortality and many have yet to implement such PLIs. The importance of timing of PLI implementation on mortality outcomes is poorly understood. We extracted cumulative daily country-specific COVID-19 mortality for France, Germany, Italy, Spain and the UK to examine country-specific mortality trends and found that all five countries experienced COVID-19 mortality epidemics initially of exponential nature. We estimated the magnitude of effect on mortality of the nationwide PLI implemented on day 18 of Italy’s mortality epidemic and assessed the importance of timing of PLI implementation by computing the effect of implementation of a PLI of this magnitude at various times on subsequent mortality. The nationwide PLI in Italy saved an estimated 6,170 lives by day 30 of the Italian epidemic. Implementing a PLI with this effect magnitude in a country of 60 million people on the day of the first death, and on days 7, 10, 14 and 17 thereafter, compared to implementation on day 18, resulted in substantially greater numbers of lives saved. Implementation on day 1 resulted in an additional 3,477 lives saved, 6,955 intensive care unit admissions and 52,162 hospital admissions prevented, beyond that achieved by implementation on day 18. PLI implementation earlier than day 18 substantially enhances benefit. Intervention on the day of the first mortality event in a country achieves the greatest benefit.

---

The global impact of COVID-19 is immense(1) and the public health threat it poses is unprecedented and immediate. Several European countries have implemented population level interventions (PLIs) at various times in their epidemics. Mathematical modelling of hypothetical PLIs predicts that early implementation of PLIs would result in substantial savings in lives lost and in burden on public health services, compared with later implementation(2). The definition of early implementation used in this modelling was a mortality rate of 0.2 per 100,000 population per week(2).

There is limited information available on the effects of real-life PLIs. The PLI in Wuhan, China was reported to significantly decrease the COVID-19 epidemic growth rate and to increase doubling time of cases(3). A very recent analysis of the effect of real-life PLIs on COVID-19 mortality in Europe estimated that across 11 countries, PLIs will have prevented 59,000 deaths up to 31^st^ March(4). The importance of the timing of implementation of real-life PLIs on mortality outcomes has not been reported, and many have suggested that implementation earlier than adopted by some European countries would result in greater benefit(5).

We therefore assessed the impact on mortality of the nationwide PLI that occurred in Italy on day 18 of its epidemic(6). We then modelled the effect achieved by that PLI, if implemented earlier than day 18 to predict the effect of earlier interventions on mortality, ICU admissions and hospital admissions. These data are of immediate and vital importance as many countries around the world are behind European countries in the timing of their own COVID-19 epidemics, and the optimal timing of implementation of PLIs for those countries is unknown, in what is an unprecedented situation.

## Methods

### Data retrieval

We chose to analyse COVID-19 deaths rather than incident cases, as deaths are not subject to variations in COVID-19 diagnostic testing practice from country to country, unlike incident cases, which are. We planned to study deaths from the date of the first death in each country, as we anticipated that these would happen around the time community spread in that country was happening extensively.

We extracted data from Worldometers (https://www.worldometers.info/coronavirus/#countries), to obtain numbers of actual reported cumulative deaths for each of the countries currently suffering major European outbreaks (Italy, Spain, France, Germany and the United Kingdom [UK]). We also extracted population numbers for each country from Worldometers (https://www.worldometers.info/world-population/population-by-country/) and used these data to calculate cumulative death rates per million head of population for each country and mortality rates per 100,000 population per week. Data were collected for each country starting on the day of the first reported death for each country, as for each of Italy, Spain, Germany and the UK, the first death was immediately followed by a rapid increase in deaths from the next day onwards, see https://www.worldometers.info/coronavirus/country/italy/ for Italy, https://www.worldometers.info/coronavirus/country/spain/ for Spain https://www.worldometers.info/coronavirus/country/germany/ for Germany and https://www.worldometers.info/coronavirus/country/uk/ for the UK, indicating the beginning of an epidemic in each of those countries. France was the exception as there were two isolated deaths (each imported to the country by a tourist) reported 16 days and 5 days before the third death, which was immediately followed by a rapid increase in deaths from the next day onwards, indicating the beginning of an epidemic for France (https://www.worldometers.info/coronavirus/country/france/). For France therefore we collected and analysed data starting on the day of the third reported death. We extracted data for Italy from the date of their first death, 21^st^ February 2020 (called day 1 for each country [except France where day 1 is the date of the third death], with each subsequent day numbered sequentially), to the last data day included in this part of our analysis (31^st^ March 2020), resulting in us having 40 days of data for Italy. The start dates and durations of epidemic data we had for each other country were as follows: France day 1 2^nd^ March 2020, duration 30 days; Spain day 1 3^rd^ March 2020, duration 29 days; UK day 1 5^th^ March 2020, duration 27 days; Germany day 1 9^th^ March 2020, duration 23 days. Dates of implementation of nationwide PLIs were obtained from(4), except that of Italy which was obtained from http://www.romatoday.it/attualita/coronavirus-roma-tutte-le-informazioni.html.

### Statistical analysis

Data were presented as estimates with 95% confidence intervals (CIs). Statistical analyses were performed using Prism 7.04 (GraphPad Software).

Each of the outbreaks were plotted individually, using cumulative death rates per million from day 1 for each country, to determine how well they fitted an exponential curve. Having observed that they fitted an exponential curve extremely closely, we then extrapolated 6 days ahead of the 31^st^ March 2020 for each country except Italy, using the GraphPad exponential curve-fit software to illustrate the exponential nature of each country’s epidemic, with 95% CIs.

The Italian outbreak was analysed to determine the magnitude of effect on mortality of the real-life PLIs implemented on day 18 during Italy’s epidemic. We examined the slopes of the natural log-transformed death rates to determine whether there were distinct phases during the epidemic, lines were fitted to these phases and trends examined using the slopes. Three phases were identified with clear changes in slope that were related in time with the localised PLI introduced in the North of Italy early in the epidemic and with the nationwide PLI implemented on day 18. However, the 95% CIs of the slope obtained for the first phase were extremely wide and this slope was therefore analysed no further. We therefore divided the entire epidemic into two phases: the initial phase of the entire epidemic from the first death to the change in slope following the nationwide PLI (day 22), and the phase thereafter from day 22 to day 30. The magnitude of the effect size of the Italian national PLI on day 30 was calculated from the slope of the latter phase, compared with the extrapolated slope of the first phase.

We then derived a formula that linked the date of a PLI to subsequent mortality rates, assuming a magnitude of effect of the PLI identical to that of the nationwide PLI in the Italian epidemic to day 30. This formula was then used to model the effect of different timings of a PLI with that effect size, upon a country with a population of 60 million people, in terms of lives saved by day 30 by PLIs implemented on day 1, day 7, day 10, day 14, day 17 and day 18 of that country’s epidemic. From, numbers of lives saved, we estimated the number of ICU admissions and hospital admissions that would also be prevented, assuming that for each death, there would be two ICU admissions, and fifteen hospital admissions, based on reported data from China, Korea and other countries(7).

## Results

### Trends in COVID-19 mortality in five European countries from the date of the first death

Trends in COVID-19 mortality from the date of the first death in each country were plotted. The first death varied from as early as 21^st^ February 2020 for Italy, to 9^th^ March 2020 for Germany, with France, Spain and the UK falling in between. All five countries displayed exponential increases in mortality from day one (the day of the first death) up to and including the last day on which data was extracted (the 31^st^ March 2020). The coefficients of fit for these exponential curves varied from 0.984 for Italy to 0.997 for Germany, showing an extremely tight fit to an exponential curve for each country and justifying our choice of day one as the true beginning of an exponential rise in mortality in each country. The tight fits of curves permitted us to extrapolate these curves to illustrate estimated mortality rates in the next 6 days for the countries other than Italy to show the exponential nature of the initial phase of each country’s epidemic. The exponential curves of actual mortality experienced to the 31^st^ March 2020 for each country are shown in Figure 1 a-e in the blue curves, and results of the extrapolations for France, Germany, Spain and the UK are shown in Figure 1 b-e in the red curves, in rates of death per million population and in days from the day of the first death, with 95% CIs shown as dotted lines.

**Figure 1.**
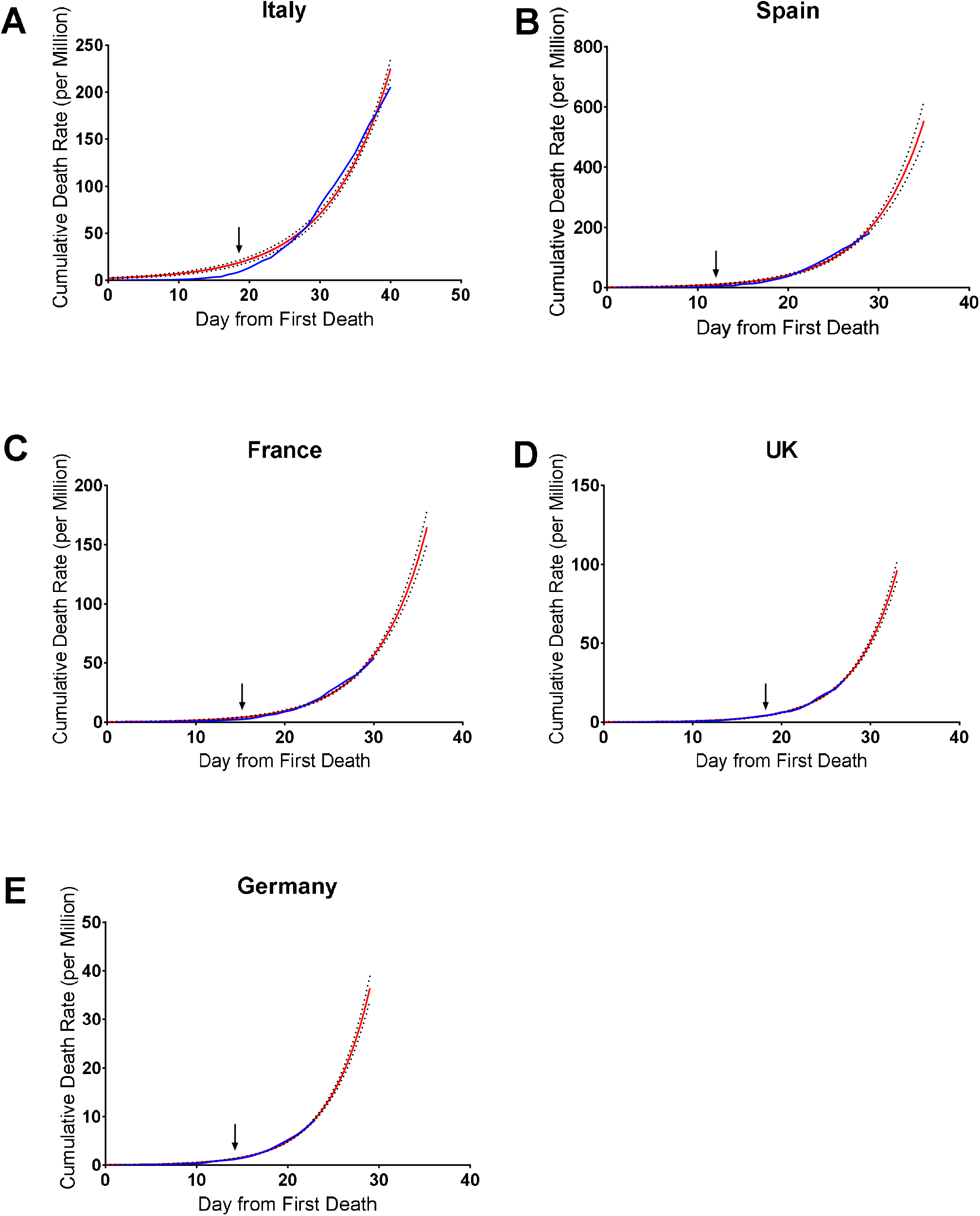
Trends in COVID-19 mortality in five European countries expressed as days from first death in rates of death per million population. A-E: death rate per million for each of the five European countries plotted against days since the first death. The blue line is the actual data to 31^st^ March 2020, with the exponential trendline shown in red. The dotted lines show the 95% CIs for the countries. All the countries but Italy are extrapolated 6 days ahead of the data for 31^st^ March 2020, in order to show the exponential nature of each country’s initial epidemic curve. The date of implementation of each country’s nationwide PLI is indicated by an arrow.

### Effect on mortality of real-life population level interventions in Italy

Italy’s mortality epidemic is the most advanced among European countries. We therefore focused our analyses on the effect of PLIs on this country. Italy undertook two PLIs which had perceptible results in terms of altering the slope of the epidemic in that country. These can be seen in Figure 2a, where there is an initial steep epidemic slope of 0.6049 (95% CI: 0.4941-0.7156) from day 1 to day 5 (blue line, Figure 2a), at which point, five days after the country introduced a regional PLI targeted at the northern Italian regions of Lombardy and Veneto, there was a change in slope to 0.2952 (95% CI: 0.2874-0.3029) (red line, Figure 2a). The second PLI was a national lockdown of the entire population, announced on the 9^th^ March (day 18 of Italy’s mortality epidemic). This was followed on day 22 by another change in slope of Italy’s epidemic slope to 0.1283 (95% CI: 0.1185-0.1381) (orange line, Figure 2a) which remained unchanged from day 22 to day 40. We confirmed that the epidemic curve from day 22 to day 30 remained exponential (R^2^ value = 0.982).

**Figure 2.**
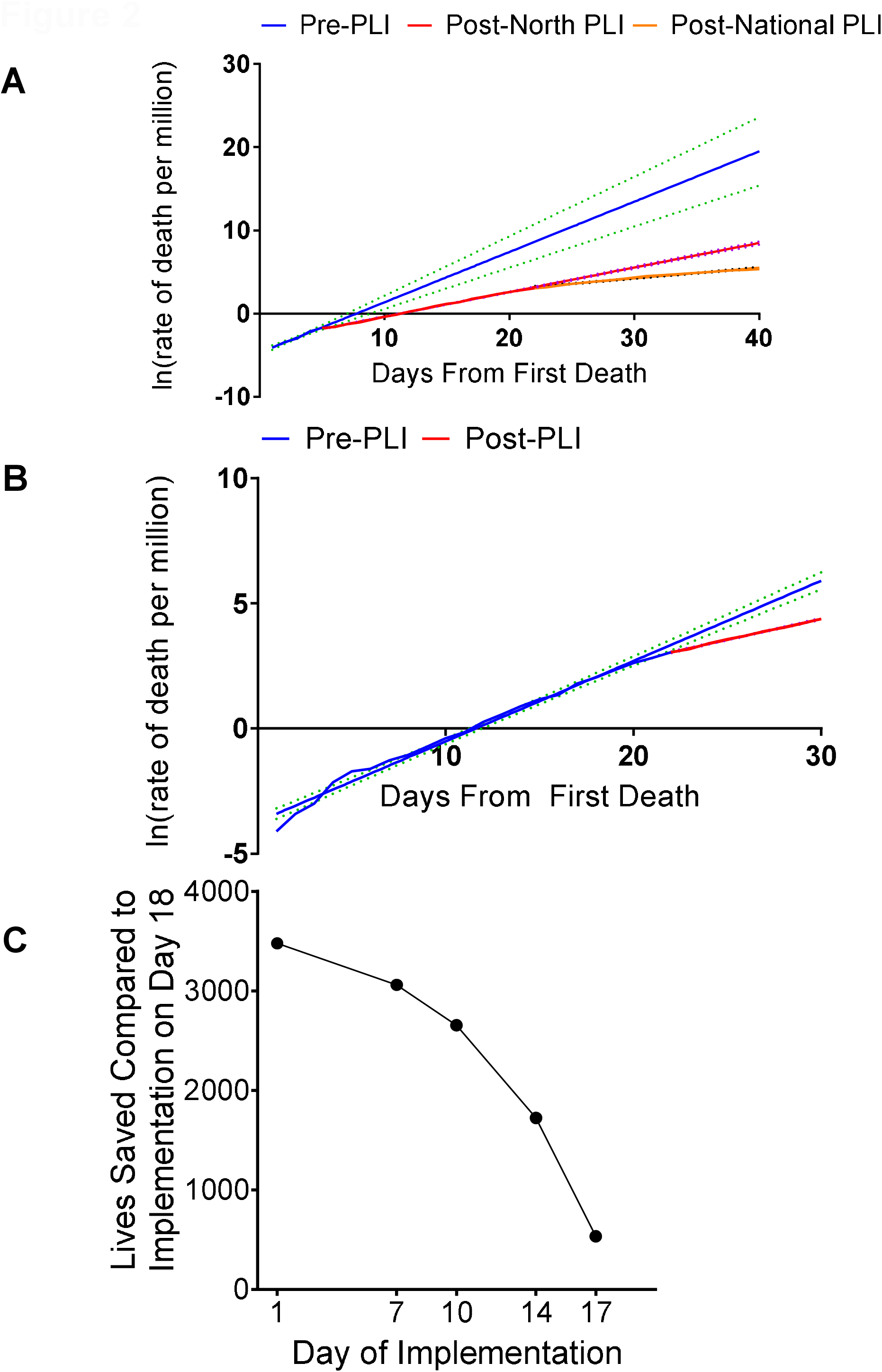
Estimation of the magnitude of the effect of PLIs in Italy and illustration of numbers of lives saved by earlier PLIs. A Three phases of the Italian epidemic were identified, by plotting the natural log transformed mortality rates against days from the first death, with the slope for each phase extrapolated to day 40 to show the magnitude of the effect of the PLIs in terms of altering the slopes of the epidemic lines. The dotted lines show the 95% CIs for the slopes of the lines. B Two phases of the Italian epidemic, shown by plotting the natural log transformed mortality rates against days from the first death. The line in blue corresponds to the average slope of the Italian epidemic before the nationwide PLI took effect, while the line in red shows the slope of the Italian epidemic after the nationwide PLI took effect. The dotted lines show the 95% CIs for the slopes of the lines. The blue line is extrapolated to Day 30 to permit comparison of Italy’s trajectory without intervention to the trajectory achieved following intervention to day 30, to permit estimation of the magnitude of effect of this PLI. C A plot of calculated numbers of lives saved compared to a PLI implemented on day 18 against the day number of implementation of earlier identical PLIs, demonstrating that the later the PLI is implemented the more dramatic the reduction in the number of lives saved.

To estimate the effect of the first PLI on mortality, we would need to extrapolate the slope before the first PLI and compare it to the extrapolated slope after the first PLI. However, the 95% CIs around the extrapolation of the slope before the first PLI are extremely wide (Figure 2a), we therefore did not analyse this change in slope any further.

In order to estimate the magnitude of the effect of the second nationwide PLI on COVID-19 mortality, we divided the entire epidemic into two phases: the initial phase of the epidemic from the first death to the day 22 change in slope following the nationwide PLI, and the phase thereafter from day 22 to day 30. As the slopes obtained for these phases had very narrow 95% CIs (Figure 2b), we used these slopes to estimate the magnitude of the effect of the PLI by examining numbers of deaths predicted by these slopes, with and without the PLI. We first examined mortality at day 30, to check whether the mortality estimated by the slope with the PLI was close to the actual mortality reported in Italy on that day (21^st^ March). This proved to be the case, as the mortality predicted by our model on day 30 (4836, Table 1) was very close indeed to the actual mortality reported in Italy on that day: 4825 deaths (https://www.worldometers.info/coronavirus/country/italy/). As the accuracy of the slopes was robust, we used them to extrapolate mortality with and without the PLI on day 30 of the Italian epidemic, and estimated numbers of lives saved by the PLI were 6,170 at day 30 (Table 1).

**Table 1.** **Magnitude of the Effect of the Nationwide PLI Implemented on Day 18 of the COVID-19 Epidemic in Italy, in terms of the Number of Lives Saved by that PLI on Day 30 of that Epidemic**.

|  | Deaths with no PLI |  | Deaths with PLI |  | Lives saved by PLI |
| --- | --- | --- | --- | --- | --- |
|  | Number | 95% CI | Number | 95% CI | Number |
| Italy (Day 30) | 11005 | (6663, 18174) | 4835 | (3249, 7196) | 6170 |

### Effect of timing of a population level intervention on mortality, ICU admissions and hospital admissions

We next examined the effect of variations in timing of implementation of a PLI. We examined only the effect of earlier interventions than the PLI implemented in Italy on day 18 of their epidemic.

We derived a formula that linked the date of a hypothetical PLI to subsequent mortality rates, assuming a magnitude of effect of the hypothetical PLI identical to that of the nationwide PLI in the Italian epidemic. This formula was then used to model the effect of different timings of PLIs with that effect size, upon country with a population of 60 million people, in terms of lives saved by PLIs implemented on day 1, day 7, day 10, day 14 and day 17 of that county’s epidemic, compared with implementation on day 18. From, numbers of lives saved, we estimated the number of ICU admissions and hospital admissions that would also be prevented, assuming that for each death, there would be two ICU admissions, and fifteen hospital admissions(7).

Assuming a 5-day time delay for effects on mortality to be seen following the implementation of a PLI (as observed for Italy) and using the steepness of the epidemic slopes calculated previously for the Italian epidemic, before and after the change in slope on day 22, following its nationwide PLI, the cumulative death rate per million on day Z was given by: ln(D) = ln(1/P) + m1(L+4) + m2(Z – L – 5), where: D=Death rate per million, P=Population in millions, m1=Epidemic slope before the change in slope following PLI, m2=Epidemic slope after the change in slope following PLI and L=Day number of PLI implementation day, counting from day 1, with day 1 as day of the first death in that country.

These analyses reveal that for a country of 60 million people, early implementation of a PLI on day 1 of such an epidemic, compared to implementation of the same PLI on day 7 of that epidemic, is calculated to result in 417 fewer deaths, 834 fewer ICU admissions and 6,252 fewer hospital admissions, by day 30 of that country’s epidemic. Comparing PLI implementation on day 1 with a later PLI implementation on day 18, as occurred in Italy, calculates early introduction on day 1 to result in 3,477 fewer deaths, 6,955 fewer ICU admissions and 52,162 fewer hospital admissions (Table 2).

**Table 2.** **Number of lives, ICU admissions and hospital admissions saved by day 30 of a COVID-19 epidemic in a country of 60 million people as a result of a PLI implemented on days earlier than day 18, compared to day 18 and compared to no PLI being implemented**.

| Day PLI implemented | Number of lives saved compared to a PLI implemented on day 18 |  | Number of lives saved compared to no PLI being implemented |  |
| --- | --- | --- | --- | --- |
|  | Number | CI | Number | CI |
| 1 | 3477 | (2614, 5415) | 10730 | (8777, 17819) |
| 7 | 3061 | (2273, 4819) | 10313 | (8436, 17224) |
| 10 | 2655 | (1955, 4215) | 9908 | (8119, 16620) |
| 14 | 1723 | (1251, 2775) | 8976 | (7414, 15179) |
| 17 | 535 | (383, 872) | 7787 | (6546, 13277) |
| 18 |  |  | 7252 | (6163, 12405) |

| Day PLI implemented | Number of ICU admissions saved compared to a PLI implemented on day 18 |  | Number of ICU admissions saved compared to no PLI being implemented |  |
| --- | --- | --- | --- | --- |
|  | Number | CI | Number | CI |
| 1 | 6955 | (5229, 10829) | 21460 | (17555, 35638) |
| 7 | 6121 | (4547, 9639) | 20626 | (16873, 34448) |
| 10 | 5311 | (3911, 8431) | 19815 | (16237, 33240) |
| 14 | 3447 | (2502, 5550) | 17951 | (14828, 30359) |
| 17 | 1069 | (766, 1745) | 15574 | (13092, 26554) |
| 18 |  |  | 14505 | (12326, 24809) |

| Day PLI implemented | Number of hospital admissions saved compared to a PLI implemented on day 18 |  | Number of hospital admissions saved compared to no PLI being implemented |  |
| --- | --- | --- | --- | --- |
|  | Number | CI | Number | CI |
| 1 | 52162 | (39215, 81220) | 160947 | (131662, 267288) |
| 7 | 45910 | (34099, 72290) | 154695 | (126546, 258358) |
| 10 | 39829 | (29331, 63230) | 148615 | (121778, 249298) |
| 14 | 25851 | (18762, 41622) | 134636 | (111210, 227690) |
| 17 | 8020 | (5743, 13085) | 116805 | (98190, 199153) |
| 18 |  |  | 108795 | (92447, 186068) |

The effect of failing to undertake a PLI at all are even more dramatic, as compared to no PLI, implementing such a PLI on day 1 is predicted to result in 10,730 lives saved, 21,460 ICU admission and 160,947 hospital admissions (Table 2).

Our calculated numbers of lives saved by Italy’s PLI implemented on day 18, and those calculated for PLI implementations on days 17, 14, 10, 7 and 1 also allow us to examine the effect of delaying implementation in terms of the magnitude of the increase in numbers of lives lost the later the PLI is implemented. We graphically illustrate this, by plotting numbers of lives saved (Figure 2c). This confirmed that leaving PLI implementation later than day 1 results in progressively increasing numbers of lives lost the longer the intervention is delayed. A similarly progressive increase in ICU (circa 2-fold greater) and hospital admissions (circa 10-15-fold greater) will also result from such delays.

### Effects on mortality, ICU and hospital admissions of timing of population level interventions in countries of varying sizes

Many countries around the world of greatly varying sizes are watching this pandemic unfold and are wondering when to respond. Table 2 is based on a linear model, and a country of 60 million people. Such countries can calculate estimates of lives, ICU admissions and hospital admissions saved, simply by dividing the numbers in Table 2 by 60 million and multiplying them by the population of their own country.

### Dates countries reached mortality rates of 0.2 per 100,000 population per week

Spain surpassed the mortality rate of 0.2 per 100,000 population per week earliest on day 17 (rate=0.228), Italy on day 21 (0.205), France on day 23 (0.203), the UK on day 25 (0.201) and Germany on day 30 (0.212). Spain implemented their national PLI 6 days earlier, Italy 4 days earlier, France 9 days earlier, the UK 8 days earlier and Germany 16 days earlier(4) than the day each country reached the mortality rate of 0.2 per 100,000 population per week.

## Discussion

France, Germany, Italy, Spain and the UK have all experienced COVID-19 mortality epidemics that were initially of an exponential nature. Each country has recently introduced nationwide PLIs at different times in their epidemic curves. The nationwide PLI in Italy implemented on day 18 of Italy’s COVID-19 epidemic, is estimated to have saved 6,170 lives by day 30 of the Italian epidemic. This is a major and very important effect, as it will have reduced ICU admissions by approximately twice as much (by circa 12,000) and hospital admissions by 10-15 times as much (by circa 60,000-90,000). It has impacted on Italy’s epidemic by reducing the steepness of its epidemic curve (Figure 1a). This change in slope is visible in the log-transformed linear plot in Figures 2a and b, on day 22 of the epidemic. COVID-19 mortality in Italy peaked on day 36 (27^th^ March 2020) at 919 deaths on that day, reducing thereafter. However, COVID-19 mortality in Italy on the 7^th^ April 2020 (day 47) was still appallingly high at over 600 deaths per day. This suggests that although this PLI was very important and produced major benefit, even greater benefit is needed. Such greater benefit might be achieved by implementing PLIs earlier.

We therefore applied data from Italy’s experience to study implementing PLIs with this effect magnitude, in a country of 60 million people and investigated whether earlier implementation would predict greater benefit. Table 2 and Figure 2c very clearly show that earlier implementation results in greater benefit with each day of earlier intervention, right back to implementation on day 1 – the day of the first death from COVID-19 in this hypothetical country. The benefits of earlier implementation are great, as a PLI implemented on day 1, compared to the same PLI implemented on day 18, was predicted to result in 3,477 fewer deaths, 6,955 fewer ICU admissions and 52,162 fewer hospital admissions. These data provide very powerful arguments for implementation of PLIs as early as possible, and we suggest that unless a death is atypical of one that is a consequence of widespread community transmission (like the first two in France, but not the first death in any other country studied), implementation on the day of the first death in any country would save a greater number of lives, ICU admissions and hospital admissions than implementation on any later day studied.

Early implementation comes with a societal and economic cost that is substantial. It is for politicians to judge the relative importance of those costs, versus the costs in terms of mortality and ICU admissions and hospital admissions. The difference between implementation on day 18 and day 1 is 3,477 deaths, 6,955 ICU admissions and 52,162 hospital admissions, indicating that (with the benefit of hindsight that those making such difficult decisions did not have at the time they had to make their decisions) day 18 is clearly too late, as hospitals will clearly be unable to cope with these numbers of ICU and hospital admissions, and almost 3,500 deaths that are clearly preventable, is clearly unacceptable.

The difference in mortality between implementation on day 1 and implementation on day 7 is 417 deaths. This translates into ∼830 ICU admissions and ∼4,000-6,000 hospital admissions. The experience of China, South Korea, Japan, Hong Kong and other Asian countries in successfully implementing PLIs to achieve control of their epidemics, but now having to maintain their PLIs long term, with relatively limited relaxations cautiously tried, indicates that PLIs in the context of COVID-19 are rather long term commitments. A difference in implementation date of just 7 days seems a small price to pay for a large effect on mortality, and even larger effects of ICU admissions and hospital admissions.

We are unfortunately not able to go back any further than the day of the first death, to determine whether implementation even earlier might bring yet greater benefits, as we have no data on mortality before this point in time, but it remains a distinct possibility that intervention before the first death has occurred might also bring greater benefits. Such earlier implementations might include regional PLIs in an area of high disease activity based on new case identification, but before a death has occurred. The dramatic change in the slope of the Italian epidemic 6 days after implementation of their regional PLI in Northern Italy (implemented on the day before Italy’s first death) strongly suggests that this regional PLI produced dramatic results. Uncertainties in the data with very wide 95% CIs, due to the relatively few data points available at that time, prevented us from estimating the magnitude of effect of this PLI with any certainty, but the magnitude of the change in slopes suggests strongly that it was very substantial.

Other countries round the world whose epidemics have started or are starting later than those in Europe, will be watching events in Europe with great interest. On the 7^th^ April 2020, there were more than 160 countries reporting less than 50 deaths (https://www.worldometers.info/coronavirus/#countries). The number of deaths reported by France, Italy, Spain and the UK on the day each country implemented its nationwide PLI were 148, 463, 196 and 281 respectively, while Germany had 94 deaths on the day it implemented. There are therefore a very large number of countries with the opportunity to implement PLIs well before they reach the numbers of deaths that had occurred at the time these five European countries implemented their nationwide PLIs. We hope the data reported herein helps to encourage those countries to implement PLIs much earlier than occurred in the European countries we have studied, with the expectation that hospital admissions, ICU admissions and mortality will be dramatically reduced as a consequence. These recommendations are supported by the data in Figure 2c, demonstrating progressively increasing numbers of lives lost the longer PLI implementation is delayed. A similarly progressive and proportionately larger increase in ICU (∼2-fold) and hospital admissions (circa 10-15-fold) will also result from such delays.

It has recently been estimated using mathematical modelling, that if a PLI is implemented early (at 0.2 deaths per 100,000 population per week) and sustained globally, then 38.7 million lives could be saved whilst if it is initiated later (1.6 deaths per 100,000 population per week) then 30.7 million lives could be saved, a difference from early to late implementation of 7 million deaths(2). We agree with the authors’ conclusion that delays in implementing PLIs will lead to worse outcomes and fewer lives saved. However, we do not feel that their adoption of a mortality rate of 0.2 deaths per 100,000 population per week for use in their mathematical modelling (no doubt based on reasonable assumptions made before the true mortality rate of this pandemic became apparent), accurately reflects early implementation in real life. Italy, Spain, the UK, France and Germany implemented their national lockdowns 4, 6, 8, 9 and 16 days respectively, before their mortality rates reached 0.2 per 100,000 per week. A rate of 0.2 deaths per 100,000 population per week in real life is therefore far too late to implement PLIs. We therefore urge countries that have not yet implemented, to do so much earlier than the rate of 0.2 deaths per 100,000 population per week used for modelling purposes, and preferably to implement before their first death if widespread community spread is already documented, or at their first death, or as soon as practically possible thereafter, if community wide information is not available.

## Data Availability

All data analysed are publicly available on https://www.worldometers.info/coronavirus/

## Author contributions

All authors contributed to the writing of the manuscript and have approved the final version for publication. Jasper Johnston and Eloïse Johnston performed the mathematic analyses and graph and table production, Sebastian Johnston lead the study design, supervision and interpretation of the studies.

